# Near real-time determination of B.1.1.7 in proportion to total SARS-CoV-2 viral load in wastewater using an allele-specific primer extension PCR strategy

**DOI:** 10.1101/2021.02.22.21252041

**Authors:** Tyson E. Graber, Élisabeth Mercier, Kamya Bhatnagar, Meghan Fuzzen, Patrick M. D’Aoust, Huy-Dung Hoang, Xin Tian, Syeda Tasneem Towhid, Julio Plaza Diaz, Tommy Alain, Ainslie Butler, Lawrence Goodridge, Mark Servos, Robert Delatolla

**Affiliations:** Children’s Hospital of Eastern Ontario Research Institute, Ottawa, Canada, K1H 8L1; Department of Civil Engineering, University of Ottawa, Ottawa, Canada, K1N 6N5; Department of Biology, University of Waterloo, Waterloo, Canada, N2L 3G1; Department of Biochemistry, Microbiology and Immunology, University of Ottawa, Ottawa, Canada, K1H 8M5; Simcoe Muskoka District Public Health Unit, Barrie, ON, Canada, L4M 6K9; Canadian Research Institute for Food Safety, Department of Food Science, University of Guelph, Guelph, ON, Canada, N1G 2W1

**Keywords:** COVID-19, alpha variant, variant of concern, WBE, PCR, public health

## Abstract

The coronavirus disease 2019 (COVID-19) pandemic caused by the severe acute respiratory syndrome coronavirus 2 (SARS-CoV-2) has claimed millions of lives to date. Antigenic drift has resulted in viral variants with putatively greater transmissibility, virulence, or both. Early and near real-time detection of these variants of concern (VOC) and the ability to accurately follow their incidence and prevalence in communities is wanting. Wastewater-based epidemiology (WBE), which uses nucleic acid amplification tests to detect viral fragments, is a faithful proxy of COVID-19 incidence and prevalence, and thus offers the potential to monitor VOC viral load in a given population. Here, we describe and validate a primer extension PCR strategy targeting a signature mutation in the N gene of SARS-CoV-2. This allows quantification of the proportional expression of B.1.1.7 versus non-B.1.1.7 alleles in wastewater without the need to employ quantitative RT-PCR standard curves. We show that the wastewater B.1.1.7 profile correlates with its clinical counterpart and benefits from a near real-time and facile data collection and reporting pipeline. This assay can be quickly implemented within a current SARS-CoV-2 WBE framework with minimal cost; allowing early and contemporaneous estimates of B.1.1.7 community transmission prior to, or in lieu of, clinical screening and identification. Our study demonstrates that this strategy can provide public health units with an additional and much needed tool to rapidly triangulate VOC incidence/prevalence with high sensitivity and lineage specificity.

## Introduction

Since the start of the COVID-19 pandemic, the possibility of SARS-CoV-2 antigenic drift, whereby genetic mutation gives rise to more transmissible and/or virulent viruses was identified as a potential risk to public health^1^. SARS-CoV-2, being a single-stranded RNA (ssRNA) virus, is susceptible to frequent mutation^2–5^. Generally, the majority of viral mutations have a low impact on disease in populations^6^, but in late 2020, several concurrent mutations emerged in circulating SARS-CoV-2 genomes which increase infectivity^7,8^ and potentially decrease sensitivity to neutralizing antibodies^9,10^. As of May 2021, these variants of concern (VOC) as declared by the WHO include: B.1.1.7 (α), B.1.351 (β), P.1 (γ), and B.1.617.2 (δ)^11^. With communities around the world already reeling from the effects of the initial pandemic waves, the putative increase in morbidity and mortality associated with VOC infection relative to that of earlier variants, is affecting the resilience of health care systems and communities.

SARS-CoV-2 wastewater-based epidemiology (WBE) is being adopted as a means of surveillance in many jurisdictions around the world (reviewed in ^12^). The common method currently used to detect fragments of SARS-CoV-2 viral RNA in wastewater is qRT-PCR utilizing probe and primer sets that were validated for clinical testing^13^. The high specificity and sensitivity of the CDC “2019-nCoV N1” (N1) and ”2019-nCoV N2” (N2) probe/primer sets seen in clinical samples^13^, appear to be mirrored in wastewater matrices^12^ although relative sensitivities between the sample types haven’t been formally investigated. N1 and N2 assays are thus used by many WBE research groups including those participating in a provincial wastewater-based surveillance initiative in Ontario, Canada. Critically, these qRT-PCR assays currently do not distinguish VOCs. Here, we describe and validate a qRT-PCR assay that targets a B.1.1.7-specific allele that lies adjacent to the N1 region and which employs new forward primers along with the existing N1 probe and reverse primer. This assay uses an allele-specific primer extension strategy, yielding excellent discrimination of single nucleotide variants in a pooled sample such as wastewater. As such, this strategy can be employed to detect other VOCs as they emerge.

## Materials and Methods

### Water resource recovery facility description and sample collection

24-hour composite samples of primary clarified sludge (PCS) were collected from the Ottawa, Ontario water resource recovery facility (WRRF) between January 16 and April 28, 2021. At initiation of sampling there was no known community transmission of B.1.1.7 in this region. 24-hour composite samples of post-grit influent and grab samples of PCS were collected from the Barrie, Ontario WRRF on January 26, 2021 during an institutional B.1.1.7 outbreak. Both sampled WRRFs are designed and operated as conventional treatment trains with influent entering screens and flowing to grit chambers to primary clarifiers to activated sludge units.

### Isolation of wastewater solids and RNA extraction

The solids in influent collected at the Barrie, Ontario WRRF were first allowed to settle at 4°C for one hour prior to decanting. Settled influent solids or PCS samples collected at Barrie or Ottawa WRRFs were well-mixed and 40 ml transferred to a 50 mL round-bottom centrifuge tube. Samples were centrifuged at 10,000 x g in a fixed angle rotor for 45 minutes at 4°C. The supernatant was discarded, taking care not to disturb the pellet. Samples were centrifuged a second time at 10,000 x g for 5 minutes and the remaining supernatant decanted. The resulting pellets were transferred to a RNase-free microfuge tube, the tared weight was recorded, and the RNA was immediately extracted. Total RNA was extracted using the RNeasy PowerMicrobiome Kit (Qiagen, Germantown, MD) on a QIAcube Connect automated extraction platform, with a modified protocol. The following changes to the manufacturer’s protocol were performed: 1) 250 ± 15 mg of sample pellet was added to the initial extraction step in place of 200 µL of liquid sample, and 2) the optional phenol-chloroform step was instead performed with Trizol LS reagent (Thermo-Fisher, Ottawa, Canada) prior to vortexing and centrifugation. The resulting aqueous phase was retained and processed as per the original manufacturer’s protocol and included the DNase treatment step. RNA was eluted in 100 µl of RNase-free water.

### Human feces collection and RNA extraction

Feces was collected from confirmed COVID-19 positive in-patients at the Ottawa Hospital. The patient specimens used in this study were anonymous donations, collected with consent. Their collection and use were approved by the Ottawa Health Science Network Research Ethics Board (OHSN-REB) under REB 20200235-01H. Approximately 200 μl of pooled patient feces was extracted using the same method as for the wastewater solids as detailed above.

### qRT-PCR

One-step qRT-PCR was performed with TaqMan^®^ Fast Virus 1-Step Master Mix (ThermoFisher, Ottawa, Canada). The N1 assay was performed using the premixed 2019-nCoV N1 Assay-RUO probe/primers set (500/125 nM, respectively) (IDT, Kanata, Canada). Allele-specific qRT-PCR was performed in parallel to detect non-B.1.1.7 alleles (D3) and the B.1.1.7 allele only (D3L) using newly designed forward primers combined with the N1 probe and reverse primers at 500, 125 and 500 nM, respectively. Sequences of the probes/primers are as follows: 5’-GACCCCAAAATCAGCGAAAT-3’ (US-CDC_2019-nCoV_N1_for); 5’-6-FAM-ACCCCGCAT/ZEN/TACGTTTGGTGGACC-IOWA BLACK FQ-3’ (US-CDC_2019-nCoV_N1_probe); 5’-TCTGGTTACTGCCAGTTGAATCTG-3’ (US-CDC_2019-nCoV_N1_rev); 5’-CATCTAAACGAACAAACTAAAATGTCTGAT-3’ (D3_for); 5’-CATCTAAACGAACAAACTAAAATGTCTCTA-3’ (D3L_for). qRT-PCR reactions were run in triplicate using 1.5 µl of RNA input in a final reaction volume of 10 μl. Cycling was performed on a CFX Connect qPCR thermocycler (Bio-Rad, Hercules, CA) as follows: RT at 50°C, 5 minutes, followed by RT inactivation, polymerase activation and template denaturation at 95°C for 20 seconds, and 45 cycles of denaturation (95°C/3s), then annealing/extension (55°C/45s). No-template controls (NTC) showed either no amplification after 45 cycles or, rarely, poor amplification above 40 cycles. Reactions were considered positive when Ct<40.

### RT-ddPCR

Singleplex one-step RT-ddPCR was performed to accurately quantify commercial standards used in generating qRT-PCR standard curves. 1-Step RT-ddPCR Advanced Kit for Probes supermix (Bio-Rad, Hercules, CA) together with CDC_N1 probe and primers (see above) were used to determine absolute copy number (cp) of N RNA in the following SARS-CoV-2 genomic RNA standards: Twist Bioscience Control 2, based on NCBI Genbank accession MN908947.3 (aka Wuhan-Hu-1, assigned to Pango lineage B) and Twist Bioscience Control 14, based on GISAID accession EPI_ISL_710528 and assigned to the B.1.1.7 (α) Pango lineage. Reactions consisted of 5 µL of serial-diluted RNA template, forward and reverse primers (working concentration of 900 nM each), probe (working concentration of 250 nM), 5 µL of supermix, 2 µL of reverse transcriptase, 1 µL of DTT, and PCR-grade water to make up the balance of the 20 µL reaction. The droplet generation, PCR reaction, quantification and data analysis was carried out as described in an earlier study^13^.

### Clinical data and human fecal samples

Clinical data included confirmed cases of COVID-19 attributed to the Public Health Unit (PHU) as determined by the Ministry of Health of Ontario’s surveillance case definition^14^. Reported cases were constrained to those residing in the PHU providing data at the time of illness. Confirmed case data was extracted from Ontario’s Public Health Case and Contact Management system (CCM), a central data depository for COVID-19 case and contact management and reporting in Ontario, managed by the Ontario Ministry of Health and accessed by PHUs. Data on VOCs is entered into this system from a data stream from the Ontario Lab Integration System (OLIS) or by manual entry by PHU staff. Designation of a confirmed case as having a variant or mutation of interest was done manually for the period of this study. Confirmed COVID-19 cases and confirmed B.1.1.7 lineage infection among people residing in the City of Barrie at the time of illness with accurate episode dates (earliest of onset of symptoms, testing date or date case reported) from January 7, 2021 to March 23, 2021 was extracted from CCM on March 25, 2021. Case data among people residing in Ottawa and testing positive between January 1, 2021 and April 28, 2021 was extracted on April 28, 2021. It is important to note that that practices related to routine screening or whole genome sequencing for VOC including B.1.1.7 varies over time in Ontario with testing practices scaling up and down in response to changing public health priorities and capacity.

### Statistical tests

Spearman’s rank correlation was determined between the proportion of B.1.1.7 wastewater alleles and clinical cases over time. A two-tailed Student’s t-test was used to determine significance of the correlation relative to the null hypothesis of zero correlation. Both were calculated using Graphpad Prism v. 9.1.2 for Mac OS.

### Proportional expression of VOC alleles

The proportional expression of D3L alleles (B.1.1.7 signal) was determined based on the equivalent primer efficiencies observed for both D3 and D3L assays in **Figure 2A, B** which allows direct comparison of Ct values that fall along the qRT-PCR standard curves derived from RNA templates. D3 and D3L Ct values were transformed into linear space and proportional expression calculated as the ratio of D3L and the sum of D3 and D3L expression. These operations are defined in the formula: 2^-Ct[D3L]^ / 2^-Ct[D3L]^ + 2^-Ct[D3]^.

## Results and Discussion

### Assay design strategy

Clinical diagnosis of a B.1.1.7 case, with its multiple mutations, requires a highly specific assay with a very high positive predictive value. This is difficult to achieve in a singleplex qRT-PCR assay, as targeting multiple, co-occurring mutations at different loci may be needed to unambiguously define a VOC. Thus, multiplexed assays or secondary screens incorporating Sanger sequencing and/or whole-genome sequencing are required to confirm the genetic lineage of a virus isolated from clinical specimens^15^. Unlike testing of individuals, wastewater samples include genomes (and fragments thereof) sourced from multiple infections and can be collected at high frequency (e.g., daily) over continuous periods. Thus, the additional longitudinal granularity in wastewater allows passive probing of individuals over the course of infection at a population level. This property provides substantial power to probe and follow the dynamics of allele frequencies in a population as infections wax and wane in an unbiased manner. We thus hypothesized that following a single, signature mutation in this context will give sufficient power to distinguish the VOC lineages. This is because other variants carrying a mutation at the same locus are likely to be rare and below detection limits as they are believed to be less transmissible and thus have a very low prevalence which does not change over time (**Figure 1A**). We found that one such signature mutation on the N gene at the amino acid level (D3L) is present in the B.1.1.7 lineage (**Figure 1B**). Moreover, a single nucleotide variant (SNV) in the wobble position of N:D3, T28282A appears to be unique to the B.1.1.7 lineage (**Figure 1C**). Serendipitously, D3L lies only 5 bp upstream from the start of the CDC 2019-nCoV N1 amplicon region, which we currently target to detect SARS-CoV-2 in wastewater^16,17^. Taking advantage of the high sensitivity of the N1 target, we designed forward primers with mutations at the 3’ end complementary to the B.1.1.7 signature allele at this locus (**Figure 1D**). This allele-specific primer extension strategy works by virtue of the inability of DNA polymerase to extend mismatched nucleotides at the 3’ end of the primer^18^. This method affords high allele specificity, while obviating the complexities and high cost associated with the development of competition probe assays. We then combined this forward primer with the existing N1 probe and reverse primer (**Figure 1D, and Materials and Methods**). We call these two new assays “D3” and “D3L” and should distinguish non-B.1.1.7 from B.1.1.7 viral lineages, respectively, when applied in the wastewater context. Note that this assay may not have a high specificity in clinical samples for viruses of the B.1.1.7 lineage as a rare mutation may occur at this location in individual cases. Partway into designing and testing this new assay, we noted the emergence of a SNV in B.1.1.7 sequences deposited at GISAID that co-occurs with the D3L mutation and is located in the middle of the D3L forward primer. This deletion mutant (A28271del) now appears to be the dominant SNV in the B.1.1.7 lineage, co-occurring with D3L at similar frequencies in the United Kingdom (**Figure 1D**). The D3L forward primer incorporates this SNV. Together, this design strategy (i.e., targeting a signature mutation that lies close to existing sensitive qPCR probe/primer sets) has the potential to create specific and sensitive qRT-PCR assays for determining proportional expression of VOCs in wastewater samples.

**Figure 1.**
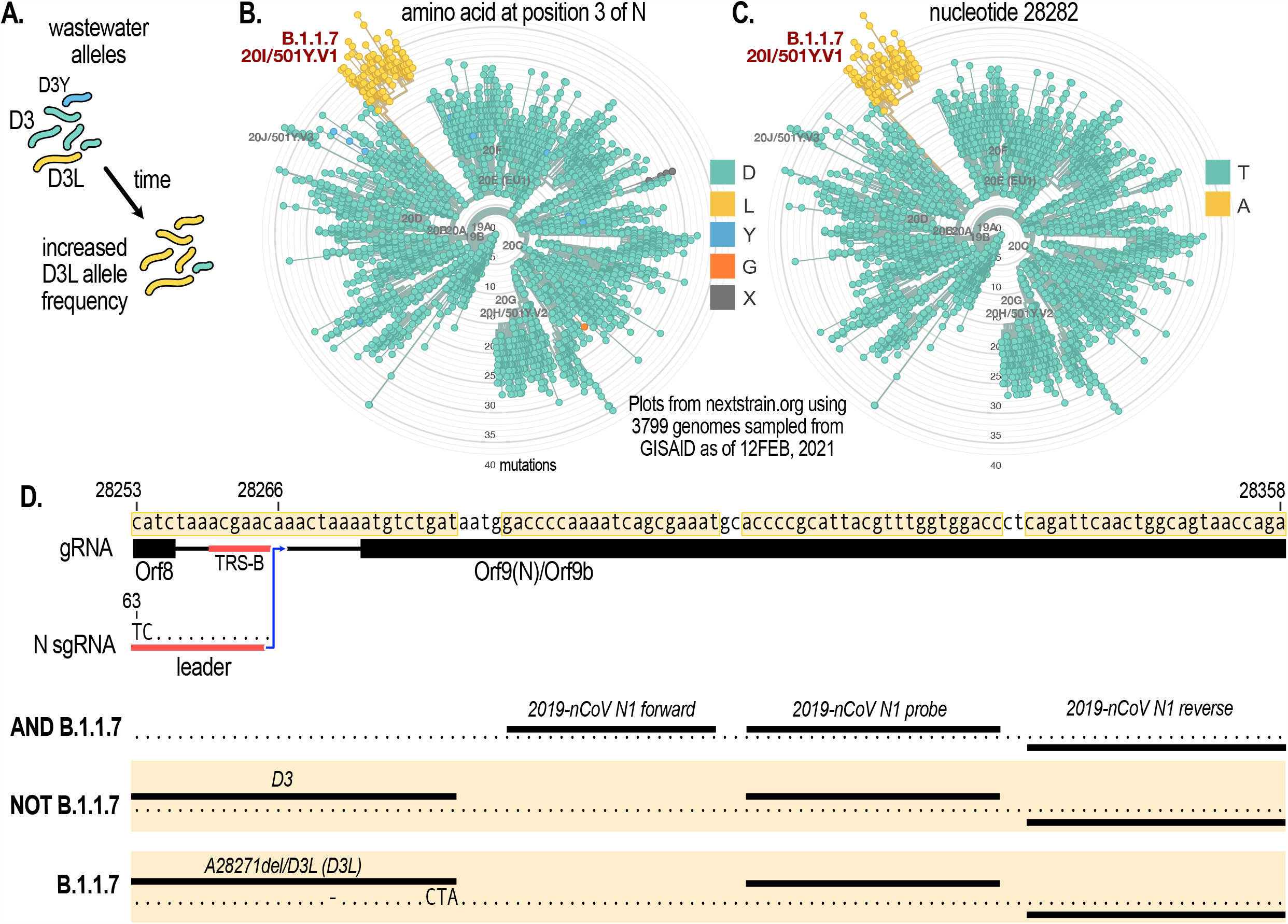
The N gene D3L mutation serves as a signature target to detect VOC of the B.1.1.7 lineage. (**A**) D3L allele frequency is hypothesized to rapidly increase in wastewater in communities where B.1.1.7 has been introduced. Other mutations at the same genomic locus which are at low prevalence would not be detected. (**B**) A radial plot of mutations amongst globally sampled GISAID SARS-CoV-2 sequences at the N:D3 locus. The D3L mutation is a signature of the B.1.1.7 lineage. (**C**) A radial plot as in **B** but at the nucleotide level showing that the T28282A SNV is unique to B.1.1.7. (**D**) Genomic neighborhood and primer/probe locations for the 3 different assays discussed in the text: N1 which amplifies 99+% of viral variants (AND B.1.1.7), D3 which amplifies alleles other than the B.1.1.7-specific allele (NOT B.1.1.7), and D3L which target B.1.1.7-specific alleles. The D3L primer also incorporates a deletion at A28271 which is present in the vast majority of B.1.1.7 genomes. TRS-B=Transcriptional regulatory site-body. Radial plots were generated on nextstrain.org^22^ with sequencing data from GISAID^23^.

### Sensitivity and allele-specificity on synthetic and patient feces-derived RNA templates

We performed D3L, D3 and N1 qRT-PCR assays on serial dilutions of B.1.1.7 or Lineage B (Wuhan-Hu-1) in vitro transcribed RNA templates obtained from a commercial supplier (Twist Bioscience, San Francisco, CA) in order to generate standard curves (**Figure 2A, B**). To compare similar template inputs, we determined copy number (cp) of the RNA templates based on N1 RT-ddPCR (indicated in **Figures 2A, B and S1**). qRT-PCR for N1 exhibited excellent efficiency (>98%) and analytical sensitivity (4/4 wells detected) down to ∼5 cp with either the B.1.1.7 or Lineage B templates (**Figure 2A, B**), consistent with our previous data^16^ and that of others^19^. Importantly, both D3L and D3 assays on their respective complementary RNA templates demonstrated excellent efficiency (98.1 and 98.4%), linearity (96 and 99%), and analytical sensitivity (4/4 wells detected) comparable to N1, although y-intercepts tended to be approximately 2 cycles higher, indicating that the limit of detection for D3 and D3L is higher than N1 under these conditions.

An allele-specific assay must be able to confidently discriminate the targeted alleles without cross-talk (i.e., it must not amplify the non-targeted allele). To this end, we assessed the D3L and D3 assays on serial dilutions of their orthogonal templates (Lineage B and B.1.1.7, respectively, **Figure 2A, B**). We observed excellent allele specificity, with no detectable cross-talk (no amplification at 45 cycles) until template copy number approached 300 (Ct∼31) at which point weak detections were observed (differences of 8-10 cycles between complementary and orthogonal templates; compare ΔCt in **Figure 2A, B**). Importantly, this cycle difference was maintained when template copies were an order of magnitude higher. To independently confirm our observations, the laboratory of Dr. Mark Servos, a participatant in the Ontario Wastewater Surveillance Initiative assessed the D3 and D3L primer efficiencies and analytical sensitivity on the same RNA templates but using a different one-step qRT-PCR supermix and found similar performance (**Figure S1**).

**Figure 2.**
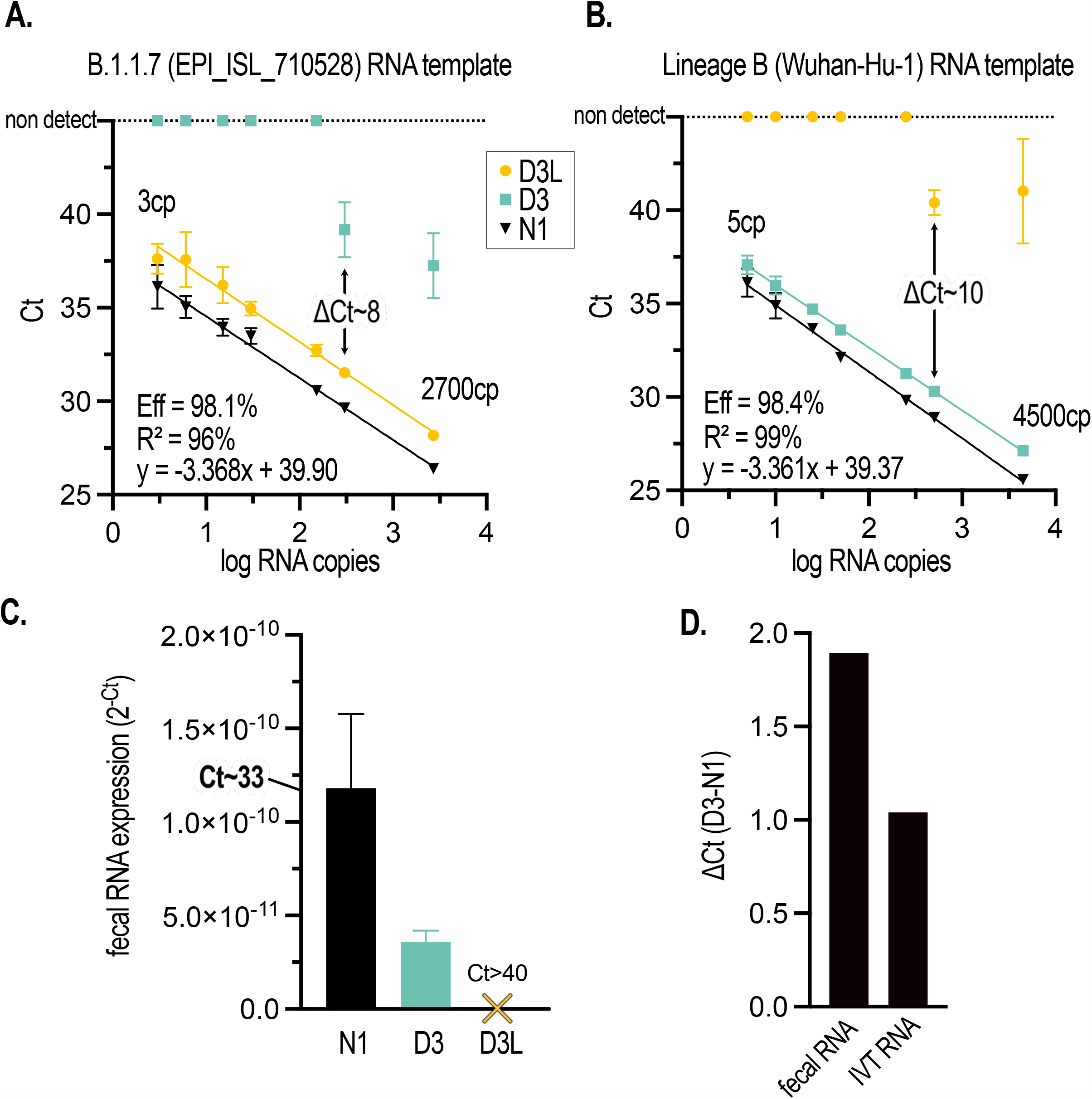
The D3L and D3 qRT-PCR assays are highly sensitive and sufficiently discriminate targeted alleles on IVT RNA templates and in COVID-19 patient fecal samples. (**A**) Standard curve of N1, D3L, and D3 assays performed with serial dilutions of a synthetic B.1.1.7 genomic RNA template (GISAID accession ID EPI_ISL_710528, Twist Biosciences). (**B**) Standard curve of N1, D3L, and D3 assays performed with serial dilutions of a synthetic non-B1.1.7 genomic RNA template (Wuhan-Hu-1 isolate). For both standard curves, N1 RNA copy number (cp) determined by RT-ddPCR is indicated. Error bars represent standard deviation of the mean of 4 qRT-PCR replicates. Calculated primer efficiencies ([10^−1/m^ -1]*100), linear fit, and equation of the line of regression are indicated for D3L (A) and D3 (B). (**C**) The B.1.1.7 allele (D3) is not detectable in RNA extracted from the feces of a human COVID-19 patient admitted to an Ottawa hospital prior to the emergence of B.1.1.7 in Canada; while both N1 and D3 (non-B.1.1.7) alleles are detected. Error bars represent the standard deviation of the mean of 2-3 qRT-PCR replicates. (**D**) D3 alleles amplify approx. 1 Ct later than the CDC N1 allele on IVT RNA, with data derived from (B); and approx. 2 Ct’s later in the context of a patient fecal matrix, with data derived from (C).

We next wished to assess the sensitivity and specificity of the D3L assay by probing fecal RNA extracted from COVID-19 hospital patients that had been admitted in Fall 2020 to the Ottawa Hospital, prior to the introduction of B.1.1.7 into the community. SARS-CoV-2 RNA isolated from human feces could be partially degraded/fragmented and contains both sub-genomic (sg) and genomic (g) RNAs. The D3L and D3 forward primers span an intergenic region of the genome and the two 5’ terminal nucleotides of the forward primers are a mismatch for the leader sequence that is present on intact N sgRNA transcripts (**Figure 1D**). Thus, it’s possible that D3L and D3 qRT-PCR sensitivity is lower in viral RNA extracted from COVID-19 patient feces compared to N1. Although we readily detected N1 amplicons (Ct∼33) in COVID-19 patient feces, we observed that D3 signal in the same sample was ∼30% of N1 (**Figure 2C**). Critically, we did not observe any amplification of the D3L allele in patient feces after 45 cycles, consistent with the allele-specificity observed with the synthetic RNA template and the putative B.1.1.7-negative status of the patients (**Figure 2C**). Using N1 as a normalization control, we compared D3 sensitivity in patient fecal RNA vs. that obtained from IVT RNA. Comparing the difference in Cts (ΔCt) between D3 and N1 signals (**Figure 2D**), we found that the D3 assay was approximately 50% less sensitive (amplified ∼1 cycle later) with a fecally-extracted vs. IVT viral RNA template. Although this data was derived from a single fecal sample, it nonetheless suggests that there is decreased binding of D3 forward primers to this genomic locus; perhaps due to increased competition from other RNAs present in the complex matrix of feces or because this region is more fragmented, or because sgRNAs are amplified less efficiently due to mismatches at the 5’ end of the forward primer. Together, these data demonstrate the sensitivity and allelic specificity of D3L and D3 assays in COVID-19 patient fecal samples.

### Application of assay to influent and primary sludge wastewaters

We obtained 24h composite influent and grab primary sludge samples collected on January 26, 2021 from a WRRF serving the mid-size (pop. ∼150K) community of Barrie, Ontario. More than two weeks prior to sampling, on January 8, a COVID-19 outbreak was declared at a long-term care facility (LTCF) within the WRRF catchment. More than 100 B.1.1.7 cases associated with this outbreak were later confirmed by whole genome sequencing of clinical samples at the provincial laboratories. The majority of cases were likely active at the time of sampling (**Figure 3A**). We readily detected N1 amplicons in both samples, indicating that SARS-CoV-2 was prevalent in the community at the time of sampling and served as a positive control (**Figure 3B**). Critically both samples from Barrie yielded detectable signal with the D3L assay (**Figure 3B**). Surprisingly, D3, targeting non-B.1.1.7 was at very low levels compared to D3L. Given that primer efficiencies of D3 and D3L are virtually identical (**Figure 2A, B)**, this allowed us to estimate their relative abundance and suggested that B.1.1.7 was the dominant viral lineage in the community at the time of sampling. To assess the sensitivity and specificity of the D3L assay in a background of non-B.1.1.7 SARS-CoV-2 in a wastewater context we sampled 24h composites of primary sludge obtained on January 16, 2021, from a wastewater resource recovery facility (WRRF) serving a large Canadian urban center (Ottawa, Ontario; pop. ∼1M). Although we could detect both N1 and D3 alleles, D3L failed to detect B.1.1.7-specific RNA fragments (**Figure 3B**). According to Ottawa Public Health, fewer than 5 clinically verified and travel-related B.1.1.7 cases were active in this community at the time of the wastewater sampling.

**Figure 3.**
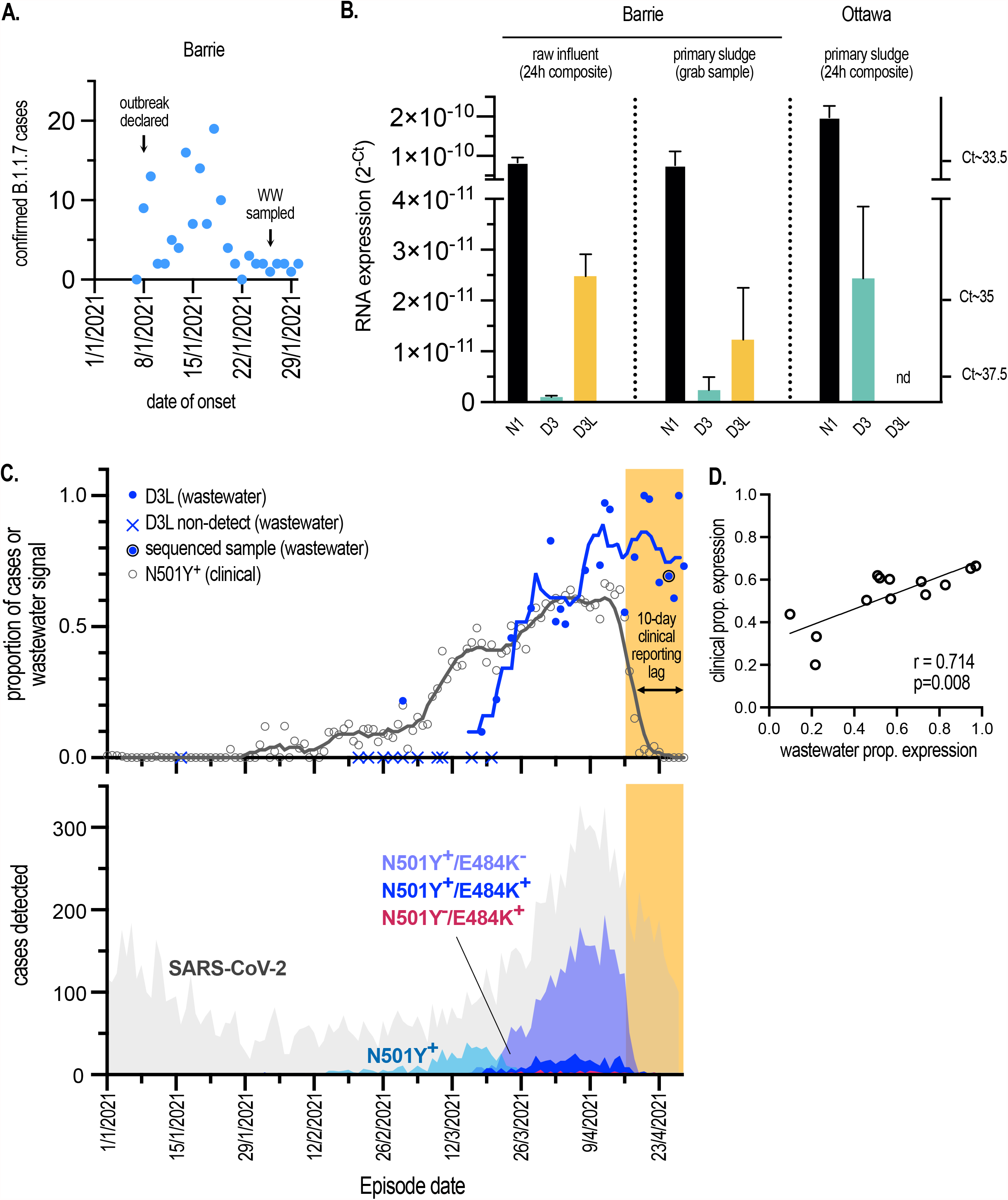
Rapid detection and quantification of B.1.1.7 proportional expression in wastewater samples following community outbreaks. (**A**) The number of B.1.1.7 cases by date of symptom onset that were associated with an isolated outbreak at a congregate living setting in Barrie, Ontario in January 2021. Cases were retrospectively assigned to the B.1.1.7 lineage by sequencing performed at Public Health Laboratories in Ontario. Wastewater (WW) was sampled from the community treatment plant serving approx. 150K inhabitants on January 26, 2021. (**B**) Expression of alleles in wastewater determined by qRT-PCR. N1, D3, D3L assays were performed on raw influent solids or primary sludge samples collected from WRRFs in i) Barrie, Ontario on January 26, 2021 and experiencing an outbreak of B.1.1.7 COVID-19 (approx. 100-200 cases), and ii) Ottawa on January 16, 2021 and with <5 known and active B.1.1.7 cases. Mean of qRT-PCR replicates (n=3) presented with error bars representing s.d. (**C, top**) The proportion of N501Y+ clinical cases from all tested cases by episode date compared to the proportional expression of D3L alleles expressed in wastewater. Trend lines represent 7-day midpoint rolling averages. “N501Y+” includes positive cases detected with the singleplex N501Y assay, as well as N501Y+/E484+ and N501Y+/E484K-genotypes assigned with the multiplex assay. Metagenomic sequencing on April 25 confirmed the major proportion of wastewater SARS-CoV-2 RNA was of B.1.1.7 origin. See text for details. (**C, bottom**) SARS-CoV-2 case distributions in Ottawa, Ontario by episode date and assigned genotypes. Cases initially screened positive by qRT-PCR (SARS-CoV-2) were subsequently screened using qRT-PCR assays targeting either N501Y beginning in February, or N501Y/E484K beginning in March. The estimated reporting lag period between the time a case is detected and the reported results of the N501Y/E484K assay is indicated by the yellow shaded region. Clinical data were provided by Ottawa Public Health and Simcoe Muskoka District Health Unit. (**D**) Spearman’s rank Correlation of wastewater D3L vs. clinical N501Y+ proportional (prop.) expression as proxies of estimation of B.1.1.7 incidence and prevalence in Ottawa. Datapoints falling in the time period between January 1, 2021 and April 15, 2021, prior to the clinical reporting lag, were included. A line of regression was computed, and Spearman’s rank correlation coefficient (r) is indicated. A two-tailed t-test indicated a significant linear relationship between the plotted variables.

To validate the D3L qRT-PCR assay, we sent the same Barrie raw influent sample to be sequenced at the National Microbiology Laboratory (NML) in Winnipeg, Manitoba. In a companion report by Landgraff et al^20^ a consensus metagenome was sequenced from this sample that was consistent with viruses of the B.1.1.7 lineage, thus confirming both our qRT-PCR data and the contemporaneous clinical landscape in the community.

### Measurement of community incidence

Passive and near-real-time monitoring tools to assess disease incidence and prevalence within communities and across larger political jurisdictions have transformative potential for public health. In the context of the current pandemic where an already highly transmissible respiratory virus has undergone antigenic drift that has led to higher attack rates, rapid intelligence and action is required to effectively curtail new outbreaks. Where the immunized predominate, breakthrough events resulting from infections with emerging viral variants must be rapidly identified. The current intensity of clinical surveillance in countries with either developing or developed health care systems is unsustainable. With strained resources, logistic weaknesses, and waning patient compliance, clinical testing will be difficult to maintain and complementary approaches highly desirable. We aimed to assess the feasibility of quantifying the proportion SARS-CoV-2 VOC in a community using the D3L/D3 assays. Twice weekly measurements for N1, D3L and D3 in Ottawa primary sludge were undertaken beginning February 21, 2021. N1 served as a VOC-agnostic positive control. We did not reliably detect (>1 of 3 replicates under 40 Ct’s) D3L alleles until March 2, 2021 at which point we computed the proportional expression of D3L alleles as a function of total alleles or signal in wastewater; a ratio of D3L (B.1.1.7) signal to total SARS-CoV-2 signal (D3+D3L). See Materials and Methods for more detail. Additional non-detects were observed until the middle of March at which point D3L allele expression consistently increased and with which we were able to calculate proportional expression (**Figure 3C, top)**. In an attempt to quantify the functional sensitivity of the D3/D3L vs. CDC N1 assays in situ (i.e., the relative ability to detect fecally shed viral RNA fragments present in the primary sludge) vs. analytical sensitivity derived in vitro (i.e., on synthetic RNA template) we measured the distribution of N1 as well as the sum of D3 and D3L signals, the latter of which is an estimate of the total signal from this genomic locus in wastewater. We observed a nearly 1 log difference in the mean of the two distributions (**Figure S2**), indicating that amplification from the D3 and D3L loci is significantly impaired in the wastewater context compared to that on synthetic RNA or template extracted from patient feces. Possible explanations include increased fragmentation in the locus targeted by the forward primers, increased competition of forward primers for other nucleic acid templates, or the fact that they overlap an intergenic region that might differentially detect genomic vs. sub-genomic RNA concentrations.

We wished to determine if the D3L proportional expression observed in wastewater correlated with VOC-specific clinical surveillance efforts. SARS-CoV-2 genomic surveillance in Ontario did not have the capacity to process the volume of cases during the Winter and Spring COVID-19 waves in Ottawa (January-April, 2021; c.f. COVID-19 cases distribution across this period in **Figure 3C, bottom**) which precluded unbiased sampling and reporting. Coincident with our assay development and in an effort to minimize sampling bias, the Public Health Ontario (PHO) laboratories developed and validated clinical qRT-PCR tests targeting the N501Y mutation shared by many of the VOCs including those of the B.1.1.7 lineage (deployed in February/March, 2021) as well as a 2^nd^ multiplexed test targeting both N501Y and E484K mutations (replacing the singleplex N501Y test in late March)^15^. Retrospective analysis of the number of clinical cases plotted as a function of presumed date of onset of symptoms or test (episode date) between January 1 and April 28, 2021 showed a rapid and sustained increase in the number of N501Y^+^ or N501Y^+^/E484K^-^ cases beginning in the middle of March and peaking by the beginning of April, consistent with the general increase in SARS-CoV-2 cases in Ottawa observed in this period (**Figure 3C**,**bottom**). Both N501Y^+^ or N501Y^+^/E484K^-^ cases were presumed to be primarily associated with B.1.1.7 lineage infection. Note the significant time-lag in clinical VOC reporting (approx.10 days) due to bottlenecks in primary and secondary screening of nasopharyngeal swabs, due to both lab and data entry delays during a period of high disease incidence in the community (personal communication, Ottawa Public Health).

We calculated the proportion of N501Y^+^ (N501Y^+^ + N501Y^+^/E484^-^ + N501Y^+^/E484K^+^; thus, capturing putative B.1.1.7 proportion) cases relative to all cases and plotted this based on episode date (**Figure 3C, top**). Remarkably, the clinical N501Y^+^ proportional expression generally followed that of the wastewater D3L proportional expression over time. Exceptions include in the first half of March when a bump in clinical proportional expression was observed that wasn’t matched in the wastewater data. We speculate that this might have been caused by sampling bias (i.e., overrepresentation of travelers or outbreaks) and non-B.1.1.7 lineages that may be captured by the N501Y or N501Y/E484K loci (including but not limited to B.1.351). Additionally, we observed a suppression of wastewater N1 signal due to snowmelt in early March that may have resulted in false negatives for D3L. We also observed a loss of correlation due to clinical reporting time-lags towards the end of the study period in late April. Notwithstanding these biases associated with both the clinical and wastewater data, when we considered the period of January 1-April 15, 2021 (omitting the clinical reporting lag period of April 16-28) we observed significant and high correlation (r=0.714, p=0.008) between clinical and wastewater proportional expression of B.1.1.7 alleles.

Finally, to again validate the reliability and specificity of the D3L qRT-PCR assay in reporting B.1.1.7 viral RNA in wastewater, an influent sampled collected at Ottawa WRRF on April 25, 2021 was sent for metagenomic sequencing to NML (timepoint indicated in **Figure 3C**) ^20^. As was seen with the Barrie sample, the Ottawa wastewater sample yielded a consensus B.1.1.7 metagenome.

## Conclusion

These data support the use of wastewater-based allele-specific qRT-PCR strategies to follow the emergence and proportional changes of VOCs in populations served by WRRFs. We demonstrate that an allele-specific qRT-PCR assay targeting a single locus on SARS-CoV-2 genomes can provide sufficiently high lineage specificity and detection sensitivity in the wastewater context. We also demonstrate good correlation between clinical data and the expression of B.1.1.7 alleles in wastewater; the latter method to monitor VOC appears less susceptible to data inaccuracies and bottlenecks associated with scaling that occurs during times of elevated disease incidence in the community. Moreover, the flexibility and economy of this proxy measure of COVID-19 incidence allows rapid and parallel implementation of multiple assays targeting other VOC-specific alleles within existing WBE surveillance architectures. In conclusion, our study demonstrates that targeting signature mutations in the N gene with allele-specific primer extension qRT-PCR represents a viable strategy to quantitatively monitor emerging VOC in wastewater over time with high sensitivity, specificity and speed.

## Data Availability

The data that support the findings of this study are available from the corresponding author upon reasonable request.

## Acknowledgements

The authors wish to acknowledge the leadership of Dr. Alex MacKenzie (CHEO-RI) in this continuing endeavour. We are grateful to Dr. Chrystal Landgraff and her team at National Microbiology Laboratory for performing the metagenomic sequencing and to Dr. Kyrylo Bessonov and Mr. Cameron McDermaid for their helpful comments in reviewing the manuscript. The authors acknowledge the crucial participation and fruitful collaborations with Water Services at the Cities of Ottawa and Barrie; public health agencies (Ottawa Public Health, Simcoe Muskoka District Public Health Units, Public Health Ontario), along with funding provided through the Ontario Wastewater Surveillance Initiative^21^. Their respective employees’ time, facilities, resources, and assistance provided throughout the study greatly contributed to this work. We gratefully acknowledge the originating and submitting laboratories responsible for sharing genetic sequence data via the GISAID Initiative, which was used in this project.

---

**Figure S1 (Related to Figure 2).**
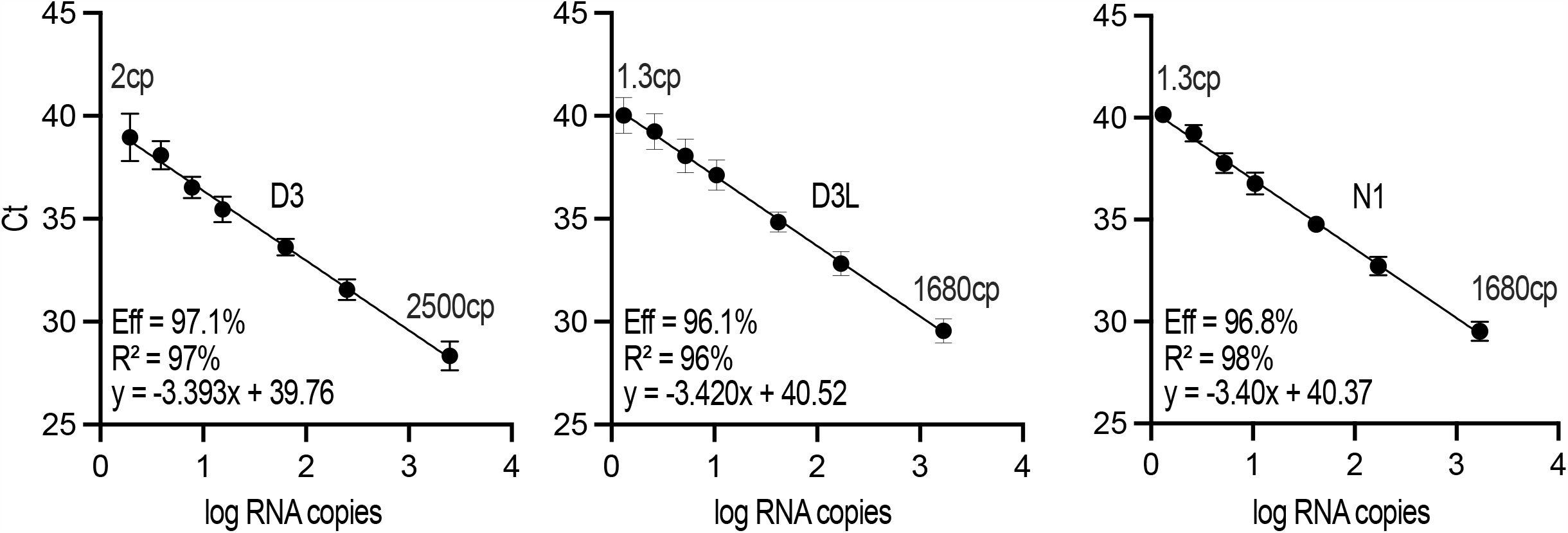
Independent confirmation of the primer efficiencies and sensitivities of the D3 and D3L qRT-PCR assays. Standard curves for D3, D3L, and N1 derived from the same templates as used in Figure 2 but using a different one-step qRT-PCR master mix were generated by a collaborating laboratory.

**Figure S2 (related to Figure 3).**
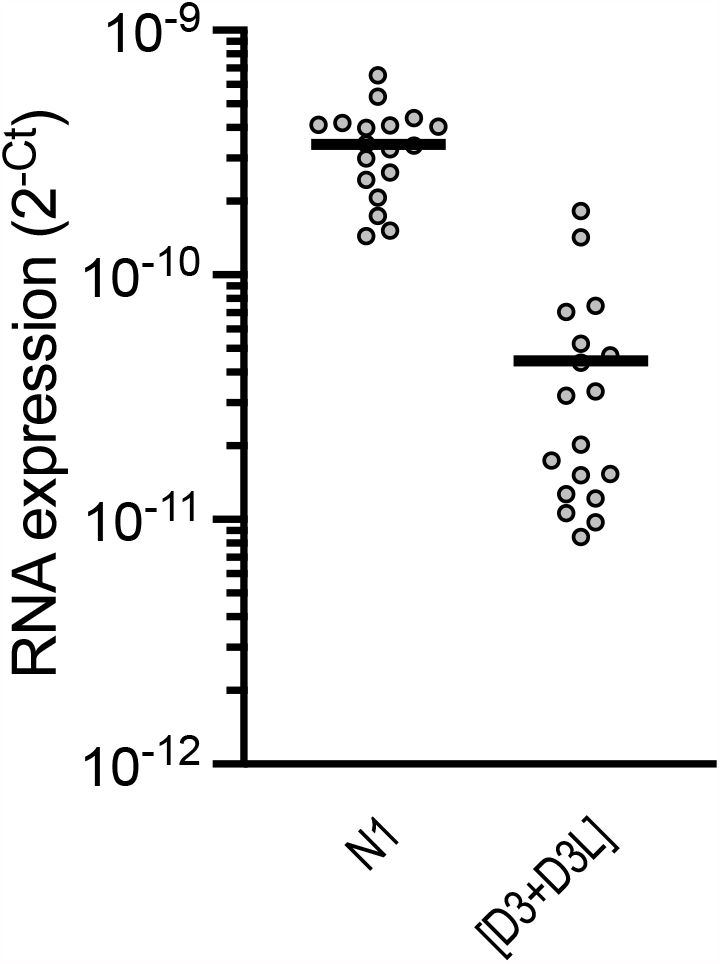
Functional sensitivity of D3L vs. N1 in wastewater. The distribution of N1 amplicon expression vs. the sum of D3 and D3L amplicon signal was compared. Both measures should represent the total signal contributed by both non-B.1.1.7 and B.1.1.7 viral RNAs but measured based on neighbouring but sufficiently different genomic loci.

